# A time series of antibiotic consumption and use at a tertiary hospital in North-western Tanzania in 2021

**DOI:** 10.1101/2025.05.15.25327629

**Authors:** S. Mapunjo, E. Magembe, E. Mayenga, J. Shao, C. Lubega, K. Makhaola, I. Lumu, the Tanzania Fleming Fund Fellowship Consortium

## Abstract

**Background:** Promoting the responsible use of antimicrobials is essential in tackling antimicrobial resistance. However, data on consumption and usage of antibiotics in Sub-Saharan Africa are still limited.

**Methods:** This was a prospective cross-sectional time series study conducted to investigate the consumption and use of third-and fourth-generation cephalosporins and fluoroquinolones in North Western Tanzania. We collect stock records from outpatient pharmacies in the hospital and conducted exist interviews each month from April to September 2021.We did descriptive analysis in Stata and Ms Excel.

**Results:** A total of 982586.2 DDD were consumed with a daily consumption of 1198.5 DDD per 1000 inhabitants per day over the six months. Five classes of antibiotics accounted for 70% of consumption. Beta lactams penicillins (J01C) are the most consumed at 329.25 DDD followed tetracyclines(J01A) 243.85 DDD. By WHO AWaRe Access antibiotics constituted 75%. Of the 253 interviews conducted 131 (51.8%) of the patients were male, 192 (75.9%) patients had bacterial infection as an indication. Ceftriaxone, was the most used cephalosporin and was used mostly to treat pneumonia while ciprofloxacin was the most used fluoroquinolone and was widely but mostly used for UTI and gastrointestinal infections. Up to 44% of prescriptions do not adhere to treatment guidelines.

**Conclusion:** We report that antibiotic consumption is in concordance WHO recommendation to have >60% of antibiotics consumed from the access group. However, there is relatively high consumption of ciprofloxacin and ceftriaxone in this hospital. Additionally, there, was significant non-adherence to treatment guidelines which underscores the need to establish functioning and robust antimicrobial stewardship programs.

**Posted history:** NONE

## INTRODUCTION

Antimicrobial resistance (AMR) represents a significant global public health concern (1,2,3). Promoting the responsible use of antimicrobials is essential in tackling AMR, as there is a clear positive correlation between the emergence of resistant bacteria and increased antibiotic consumption (4). Additionally, numerous studies have reported a consistent rise in antimicrobial drug consumption over time (4,5). Hence, surveillance of antimicrobial consumption (AMC) is important for tracking usage trends, pinpointing areas for improvement in prescribing practices, predict potential emergence of resistant pathogens, and guide targeted interventions to mitigate the growing threat of AMR

The World Health Organisation (WHO) Antimicrobial consumption defined as quantities of antimicrobials utilized in a specific setting during a specific period and is a proxy estimate for antibiotic use (6). The WHO has classified third- and fourth-generation cephalosporin along with fluoroquinolones, as critically important antibiotics. This designation is based on their role as the treatment options for certain serious human bacterial infections (7). Additionally, these antibiotics are used to treat diseases caused by organisms that can be transmitted from non-human sources to humans as well as human diseases linked to pathogens capable of acquiring resistance genes from non-human sources (8). The WHO AWaRe (Access, Watch, and Reserve) places these third-and fourth-generation fluoroquinolones and cephalosporins in the Watch group. Watch antibiotics generally have a higher potential for the selection of antimicrobial resistance and are more commonly used in sicker patients in the hospital facility setting (9).

The irrational use of antibiotics is a major factor fueling the development of antimicrobial resistance. Irrational use of antibiotics includes prescription without laboratory evidence, polypharmacy, non-prescription sales, and prescription of incorrect doses, self-medication and treatment of non-bacterial illness (10,11). The widespread and frequent improper use of fluoroquinolones has significantly contributed to the alarming increase in quinolone-resistant E. coli, resulting in greater dependence on third-generation cephalosporins as alternative treatment options in some countries (12). In Tanzania, studies have reported a high prevalence of antibiotic use, up 60% of patients in primary health care received antibiotics (13), and approximately one-third of all clients buying antibiotics from drug outlets do not have a prescription (14).

However, there are limited studies to assess the consumption of antibiotics for outpatients in a tertiary hospital setting. Therefore, this study aimed to assess the consumption and use of third-and fourth-generation cephalosporins and fluoroquinolones relative to other systemic antibacterial agents in a tertiary hospital in North Western Tanzania.

## METHODS

A prospective cross-sectional time series study conducted to investigate the consumption and use of third-and fourth-generation cephalosporins along with fluoroquinolones in a zonal hospital in north western Tanzania from April to September 2021. The study centre is a 950-bed tertiary referral hospital, serving over 14 million people across eight regions.

### Antibiotic consumption

Within the hospital, pharmacies are categorized by location and the type of clients served. The National Health Insurance (NHIF) Pharmacy for NHIF clients, F4 Pharmacy for surgical and OPD clients; and D6 and D8 Pharmacies for other inpatients. Although, a Main Pharmacy exists, collecting data from the five sub-Pharmacies would point a more accurate picture on consumption. Electronic stock records of antibiotics issued to the five pharmacies were collected at the end of every month to determine their consumptions.

### Antibiotic use

We conducted exit interviews with patients from the pharmacy window on 3-5 consecutive days of each month from April 2021 through September 2021. Based on previous studies we estimated that 150 interviews were sufficient. However, we interviewed 253 out-patients and purposively targeted patients prescribed fluoroquinolone, and generation cephalosporin. We collected data on age, sex, diagnosis, antibiotic type, dosage, route of administration and duration. We calculated the proportion of prescriptions which complied with the Standard Treatment Guidelines (STG/NEMLIT). Adherence to guidelines was defined as a composite variable (right antibiotic regimen, dosage, frequency, route of administration and treatment duration). During the study, Standard Treatment Guidelines (STGs) were up date, the major change was the replacement of ciprofloxacin with nitrofurantoin as treatment of choice for uncomplicated UTI (19,20).

### Data management and statistical analysis

The WHO Anatomical Therapeutic Chemical (ATC) classification system was used to classify antibiotic into standard codes. The amount of antibiotic consumed was expressed as Daily Defined Dose (DDD)based on the WHO guide. The DDD was adjusted for an outpatient setting and presented as DDD per 1000 inhabitants (6). The total number of patients that attended the hospital for that period was used a denominator. This data on attendance was obtained from the hospital information management system. The WHO AwaRe categorization was used to group antibiotics into Access, Aware and Reserve groups (6), the amount antibiotic was expressed in DDD.

The data was cleaned in Ms Excel and then entered into the AMC Tool version 2019 https://amu-tools.org/amctool/amctool.html. The tool has standard DDD metric for each antibiotic. The unit for each antibiotic was defined as the smallest unit form of the antibiotic for example tablets, capsules, ampoules or vials.

Key metrics derived included the total consumption of systemic antibiotics (ATC group J01) measured as DDD per 1000 inhabitants per day, their relative consumption in percentages, trends over time, concordance with the WHO AWaRe recommendations, adherence to the WHO global indicator (which recommends 60% of total consumption as Access agents), and the composition of the top 75% of drug utilization (DU75%). Antibiotic consumptions are presented by months, class, and the World Health Organization (WHO) AWaRe classification. Descriptive analysis was done using frequency and percentages for categorical variables, while medians or means were used for continuous variables. We ranked antibiotic cumulatively to the number that contributed to 70% of consumption. Analysis was done Microsoft Excel and Stata version 15.

### Ethical Consideration

Ethical clearance to conduct this study was issued by the National Institute for Medical Research with Reference code NIMR/HQ/R.8a/Vol.IX/3554.

## RESULTS

### Antimicrobial Consumption

During the six months period at total of 982586.2 DDD were consumed. However, the daily consumption was 1198.5 DDD per 1000 inhabitant per day (DDD/1000inh/day). Just 5 classes of antibiotics contributed to 70% of consumption. By ATC, Beta lactams penicillins (J01C) are the most consumed at 329.25 DDD followed by tetracyclines(J01A) 121.9 DDD while the consumption of quinolones (J01M) was 100.2 DDD (figure 1). By sub class, tetracyclines, combined penicillin, Nitrofuran derivatives, macrolides, fluoroquinolones accounted for 70% of the antibiotic consumed. The daily consumption of most antibiotics relatively followed a similar trend throughout the 6 months period spiking in June followed by a dip in July. However, it raised again in August and remained stable until September. A further break down to class level revealed that ciprofloxacin was the most consumed compared to other antibiotics and its consumption increased from April peaking in June at 24.21 DDD/1000 inh/day and it dipped in July followed by a rise reaching 22.87 DDD/1000inh/day in September (figure 2). Ceftriaxone was the most consumed Cephalosporine and consumption remained largely stable through the six months period although it peaked to 12.18 DDD/1000/inh in June. Meanwhile cefixime consumption remain stable at <1 DDD/1000inh/day (figure 2). The consumptions of fluoroquinolones were proportion higher in comparison to other third and fourth generation cephalosporin each month of the study (*figure 2*).

**Figure 1:**
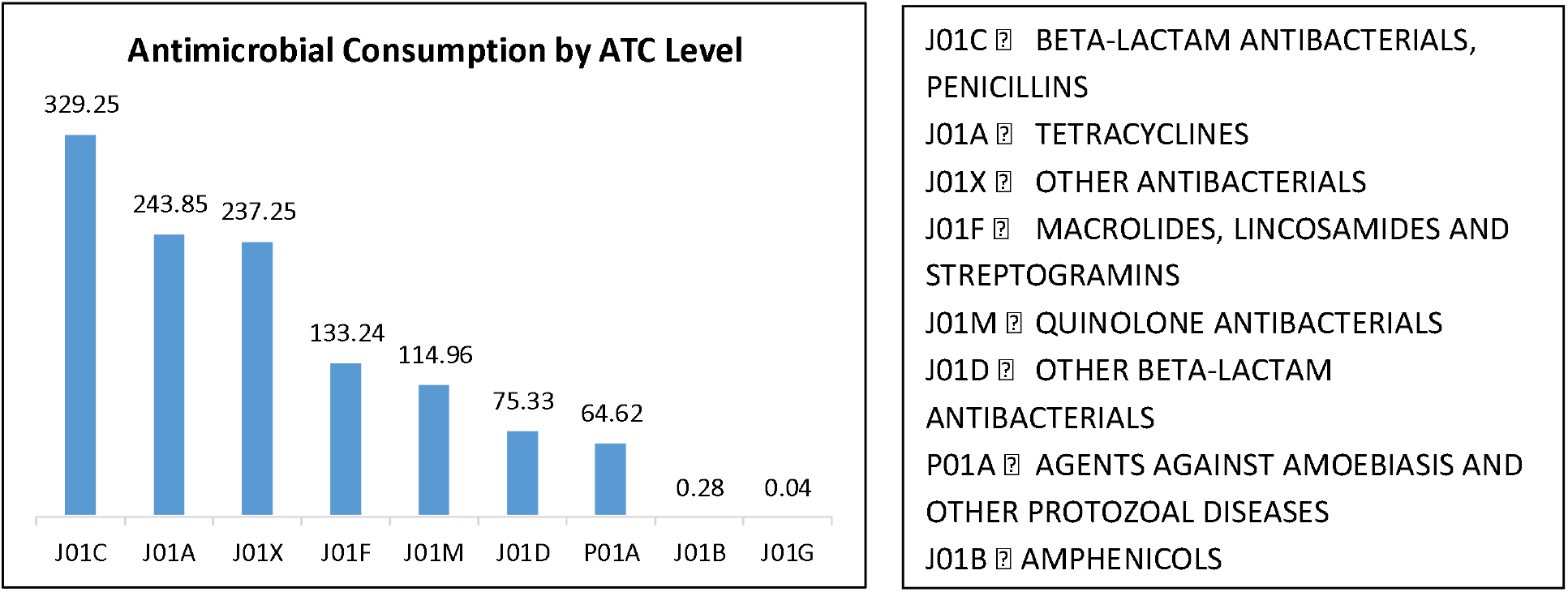
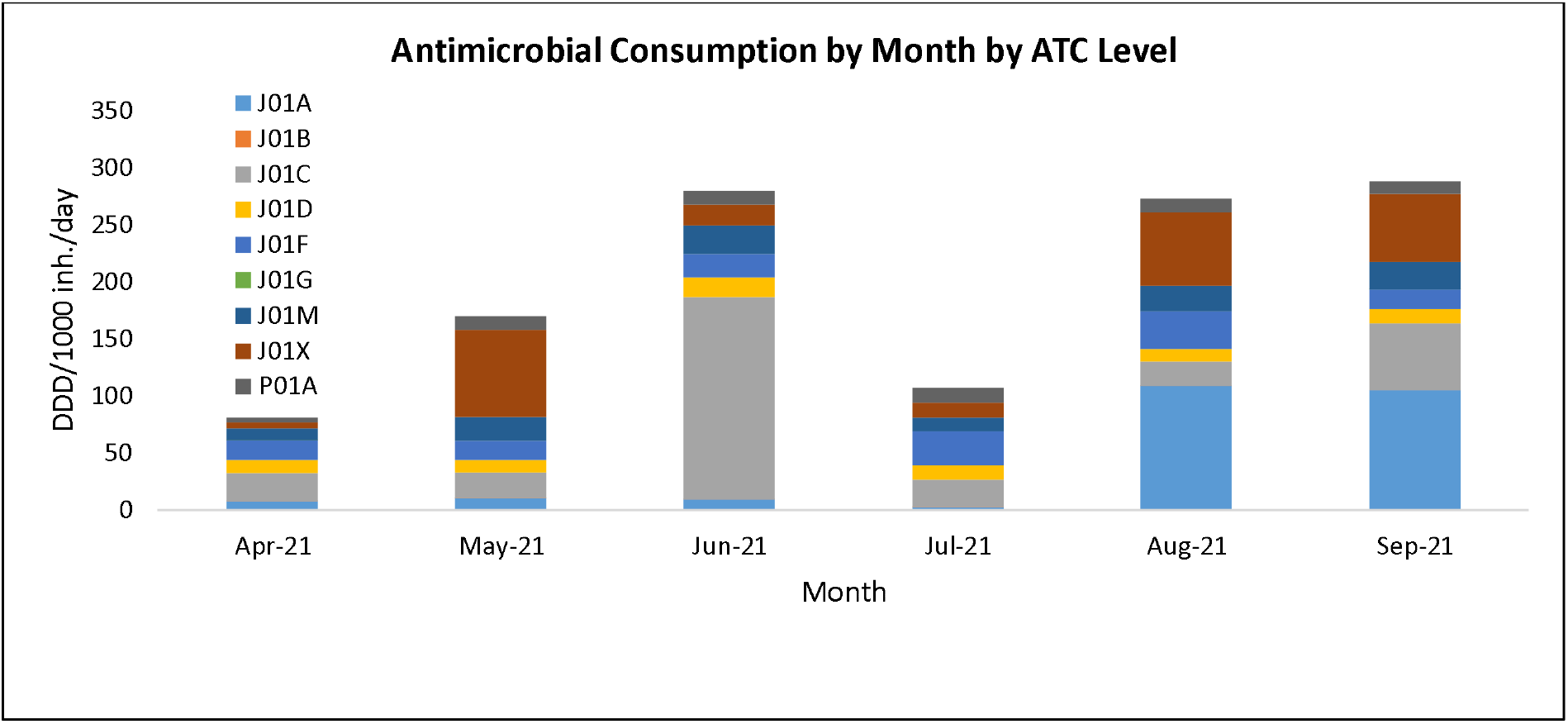
Antimicrobial consumption by ATC and month

**Figure 2:**
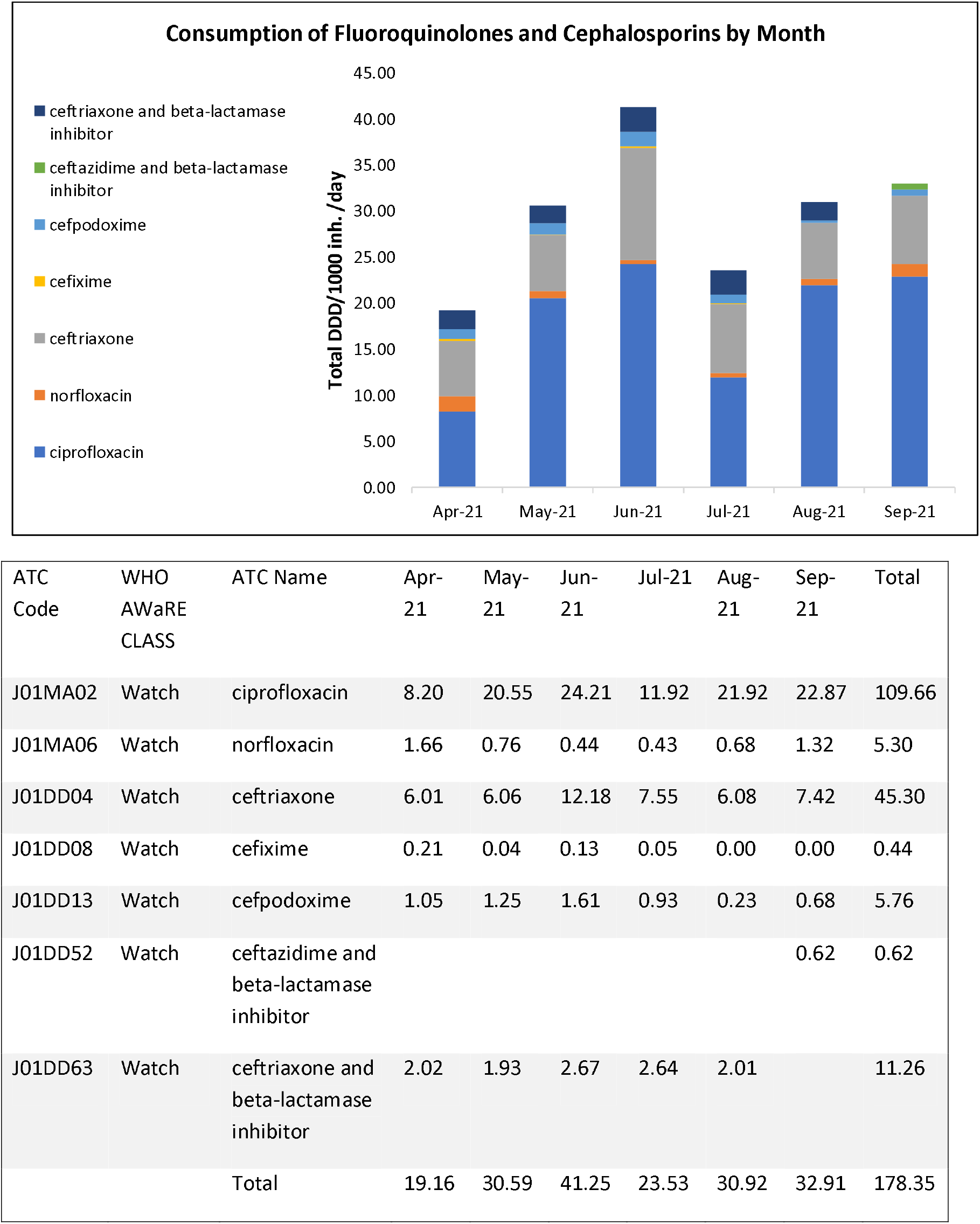
Consumption of fluoroquinolones and cephalosporin by month

Overall, 74% of antibiotics consumption were Access, 25% Watch, 1% Reserve/unclassified (supplementary figure 1).

### Antibiotic Use

During the six months period, a total of 253 exit interviews were conducted, the mean age was 46.2 (SD 23.4), and 131(51.8%) were male. Up to 192 (75.9%) patients had an indication of the prescription form (Table 1).

**Table 1:**
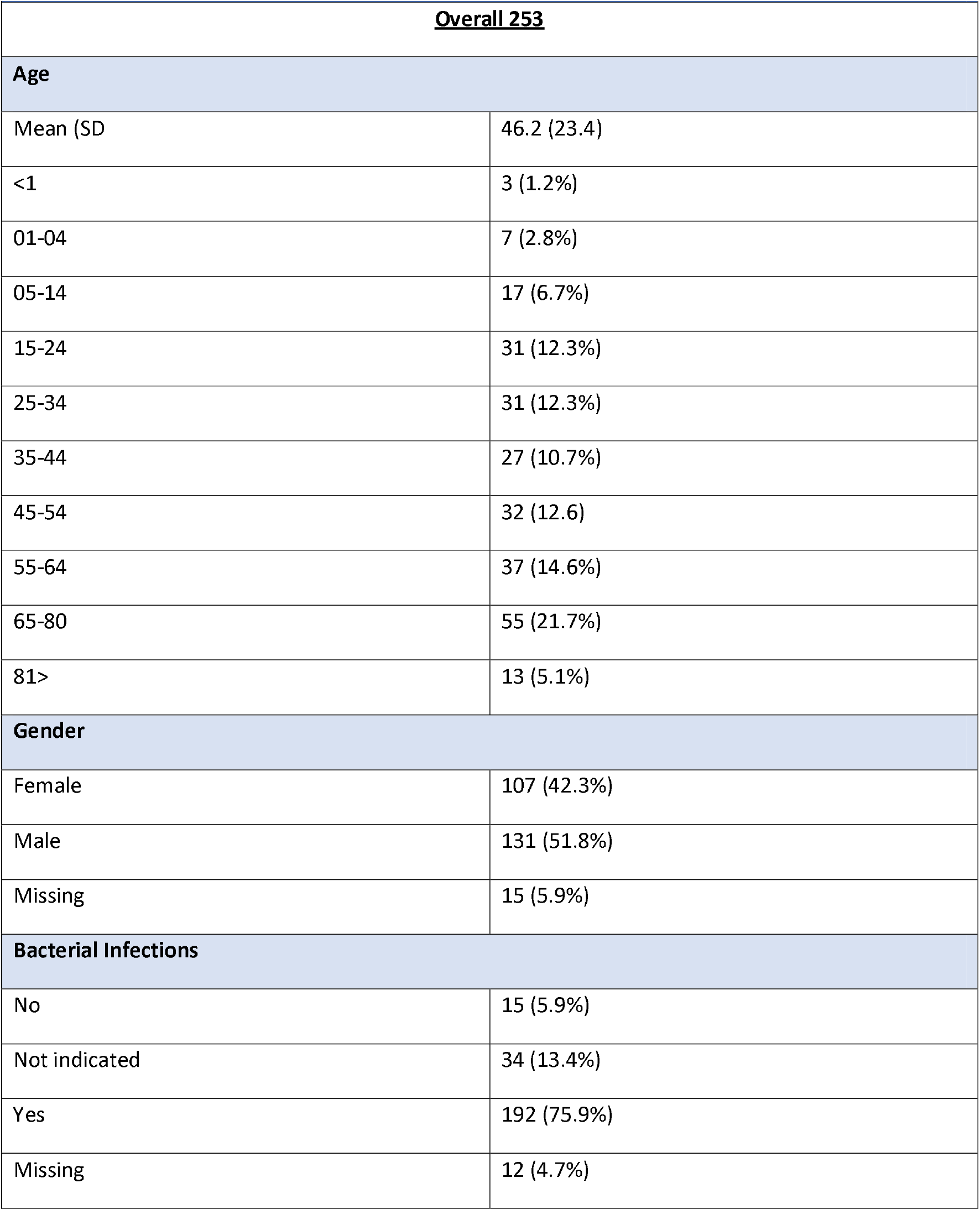
Demographic and Clinical Characteristics of Patients Sampled.

The top five indication for antibiotic were urinary tract infections, septicemia, pneumonia., urethritis and pelvic inflammatory disease (PID). Meaning these were the most common indication for use of third and fourth generation fluoroquinolones and cephalosporins (Figure 2). The top five frequently prescribed antibiotics for these conditions were ciprofloxacin, Nitrofuratoin, ceftriaxone salbactum, ceftriaxone, cefixime. Patients with pneumonia frequently received, cefixime or ceftriaxone. However, ceftriaxone, had various uses (Figure, 3). Ciprofloxacin was used for UTIs, gastrointestinal (GI) infections and but mostly for ‘other infections’, while ceftriaxone + sulbactam was the most frequently prescribed antibiotic for septicemia diagnosis (figure 3). The number of prescriptions for each antibiotic varied by month; however, ciprofloxacin prescriptions were relatively stable through the six months, while ceftriaxone/salbactum was used from July to September (supplementary figure 2).

**Figure 3:**
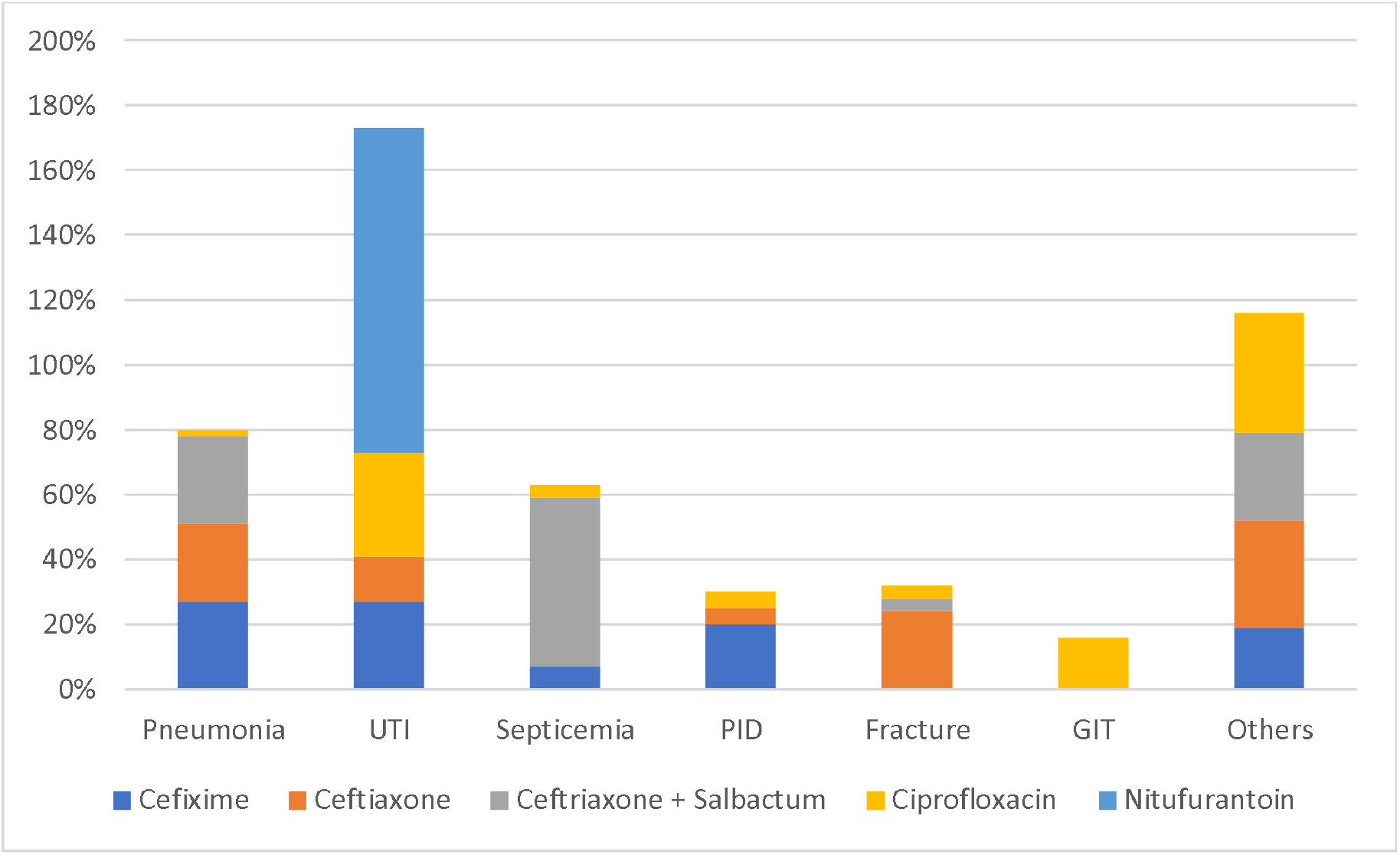
Treatment regimen for most diagnoses

Further examination of appropriate of use revealed that 56% of antibiotic prescribed adhered to the local treatment guidelines with regard to antibiotic regimen, dosage, frequency, route of administration and treatment duration while 44% had one or of the items of the composite variable wrong. On average antibiotics were prescribed for 7 days.

## DISCUSSION

In this study, beta lactam, tetracycline, and quinolones were the most consumed antibiotics. However, the consumption of the medicine varied over the six months period which could due seasonal demand exacerbated by the COVID-19 pandemic. Also, this variation may be due to challenges with stock management. Overall, the consumption of 3^rd^ and 4^th^ generation cephalosporines was significantly low compared to penicillins constituting only 5% of all antibiotics. However, an increase in the use of cephalosporines been reported in Europe and globally (16) and another retrospective study from Asia that evaluated antibiotic prescriptions in a pediatric ward revealed that, 88.1% prescriptions contained cephalosporin’s, among which 75% were 3^rd^ generations cephalosporins (17).

We observed high consumption of ciprofloxacin with a total of 45,142 DDD contributing to 95.4% of all fluoroquinolones consumed in six months. Fluoroquinolone accounted for 17% of the consumption. A high utilization of fluoroquinolones was reported in one study that evaluated use of antimicrobial in 53 countries indicated that, fluoroquinolones were among the top three worldwide (16). We found, ciprofloxacin was used widely in various conditions besides treatment of UTIs (28.5%), PID (4.6%), typhoid fever (3.8%) and septicemia (3.8%). Another study reported similar partners, ciprofloxacin was used for treatment of many disease conditions including UTI (30.8%), gastrointestinal infections such as typhoid fever (10%), PID (7%) and respiratory infections (7%) (18). The large consumption and varied used of ciprofloxacin would be driven by affordability, ease of dosing and available of ciprofloxacin as an oral tablet. Other classes of flouroquinolons for example levofloxacin are now reserved for the treatment of multidrug resistant TB.

At this zonal hospital nearly 74% of antibiotic consumed were in the access category which is in concordance with the WHO recommendations, which advocate for at least 60% of antibiotic consumption to come from access group antibiotics. Adherence to this standard not only drives the cost health care down but also prevent unnecessary consumption of antibiotics reserved for difficult to treat infections. This observation is similar to another retrospective study conducted by the Mapping of antimicrobial resistance and consumption project conducted between 2016-2018 and reported antibiotic consumption at hospital level at 68.3% in Tanzania (19). However, this proportion is lower than was previously reported in national retrospective study based on import data, where access was >90% (20). Although the consumption of more access is recommended, the consumption of limited class of antibiotics may increase selection pressure and select for particular resistance trends (19).

Among cephalosporines, ceftriaxone was the most widely consumed 3^rd^ generation cephalosporin. It was mostly prescribed for treatment of bacterial pneumonia (24%), UTI (14%), and PID (5%). One observations study among in-patients found a high prevalence of ceftriaxone use especially during surgical prophylaxis (21). The frequent use of ceftriaxone, would due to is affordability ease of administration and especially as a once daily dose. Other consumed third generation cephalosporines were cefixime (0.7%), cefpodoxime (9.2%) and ceftriaxone with beta-lactamase inhibitor at (18.7%). Ceftriaxone-Sulbactam was widely used in the management of septicemia and pneumonia. The most frequently prescribed ceftriaxone regimen was a dose of 1g (91%), administered once daily (95.5%), with an average treatment duration of 4.5 days (range 1-7 days). Given that these are out-patient pharmacies, we are not sure why, intravenous (IV) antibiotics were stocked and dispensed in these pharmacies, the assumption must these were prescribed partly for Outpatients parental Antimicrobial Therapy (OPAT). For example, in the treatment of PID or gonorrhea where it is prescribed as a start dose of 1g intramuscularly.

We found a low adherence 56% to STG/NEMLIT when these antibiotics were dispensed. In some case, the indication for treatment was not started in other cases the duration, route, frequency or dose was not appropriate. The wide spread inappropriate use of antibiotics has been widely reported in various studies across Africa sub-Saharan Africa. For example, a retrospective study in south Africa found that adherence to guideline in primary care facilities was at 45.1% (24). Another conducted in Dodoma Tanzania, revealed that only 29.9% of prescriptions in primary care adhered to the national Standard Treatment Guidelines (25). The low adherence to guideline may also be explained by the mismatch between national guidelines and the complexity of the cases seen at this tertiary hospital. The observed inappropriate use would be due to the absence of functional antimicrobial stewardship in the hospital at the time this study was conducted.

Our study provides baseline information on the trend of consumption of fluoroquinolones, third and fourth generation cephalosporines in northern Tanzania. Although, this was prospective study, the observation time was short to make meaningful interpretation of the trends. Moreover, only medicines with an assigned ATC code and DDD are included in the analyses. For example, flucloxacillin + amoxicillin was highly prescribed at this tertiary hospital, however, there is no ATC for a combined formulation hence the DDD was derived separately flucloxacillin and amoxicillin. Secondly, there was limited information on the diagnosis. For example, the patient who were prescribed antibiotic for malignance or fracture, there were no details on clear details on the diagnosis and the reason for using these antibiotics. In such case, the assumption was made that antibiotic was used unnecessary. Additionally, the medical records did not specify whether the urinary tract infection was considered complicated, uncomplicated, catheter associated or acute pyelonephritis which is crucial information for determining the appropriate treatment plan.

In summary, three quarters of antibiotic consumed are access which is in concordance to WHO recommendation to have >60% of antibiotics consumed from the Access category. However, there is relatively high consumption ciprofloxacin and ceftriaxone at the hospital. Additionally, there low adherence to treatment guideline at this tertiary hospital which underscore the urgent need for antibiotic stewardship programs at the hospital. Further studies are neeed link the trends in consumption and use of these antibiotics antibiotic resistance trend in this population.

## Data Availability

The data is available via reasonable requests directed to the Tanzania Fleming Fund Fellowship consortium.

## Contribution of the of the authors

MS, LI: conceptualised and designed the study and drafted the first manuscript.

LC LI: Analysed and interpreted the data.

MS, Shoa, LI: Wrote the manuscript

LI, KM: supervised the writing of the manuscript and critically reviewed the manuscript

We thank the study staff at Bugando Medical centre,Deus, Sinibagiye, Gabriel Maganga and John Pemba the chief pharmacist at BMC for the guidance.

## Involvement of patients

Moving forward we intent to share results of the study with patients’ committees at the hospital and encourage them to discuss the indication for antibiotics with their physicians. We hope that this discussion will in-turn lead to more rational use of antibiotics in the hospital.

## The Tanzania Fleming Fund Fellowship Consortium

Nyambura Moremi., Kolader Marion., Schultsz Constance., Shumba.Edwin., Ondoa Pascale., Richard Walemwa., Beverly Egyir., Neema Kamala., Augustin Malinga, Kayuni Gibbonce., Oskam Linda.

