## supplementary material for "A time series of antibiotic consumption and use at a tertiary hospital in North-western Tanzania in 2021"

***Figure 1: Antimicrobial Consumption by WHO AWaRE Classification***

Antimicrobial Consumption by WHO AWaRE Classification

|  |  |
| --- | --- |
| Access | 886.664 |
| Reserve and Unclassified | 11.8846 |
| Watch | 300.289 |

***Supplementary figure2: prescriptions of different antibiotics sub-classes by month***

Antibiotic Prescriptions by Month

|  | Cefixime | Ceftriaxone | Ceftriaxone + salbactum | Cephalexin | Ciprofloxacin | Norfloxacin | Nitrofurantoin |
| --- | --- | --- | --- | --- | --- | --- | --- |
| 2021-04-01 | 12 | 5 |  |  | 12 |  | 11 |
| 2021-05-01 | 3 | 5 |  | 4 | 22 |  | 7 |
| 2021-06-01 | 1 | 8 |  |  | 21 | 4 | 6 |
| 2021-07-01 |  | 4 | 2 |  | 26 |  | 8 |
| 2021-08-01 |  |  | 10 |  | 25 |  | 5 |
| 2021-09-01 |  |  | 14 |  | 25 |  | 13 |

**Supplementary table 1 Consumption by Antibiotic Class by Month (Total DDD/1000 inh. /day)**

| ATC CODE | ATC Name | Apr-21 | May-21 | Jun-21 | Jul-21 | Aug-21 | Sep-21 | Total |
| --- | --- | --- | --- | --- | --- | --- | --- | --- |
| J01AA | Tetracyclines | 8.08 | 10.46 | 9.33 | 2.61 | 108.52 | 104.84 | 243.85 |
| J01CR | Combinations of penicillins, incl. beta-lactamase inhibitors | 10.00 | 10.38 | 108.28 | 9.72 | 8.49 | 17.08 | 163.95 |
| J01XE | Nitrofuran derivatives | 3.43 | 5.74 | 3.99 | 3.98 | 58.93 | 55.38 | 131.45 |
| J01FA | Macrolides | 17.21 | 16.31 | 20.24 | 29.53 | 32.10 | 15.06 | 130.46 |
| J01MA | Fluoroquinolones | 9.86 | 21.31 | 24.65 | 12.35 | 22.60 | 24.19 | 114.96 |
| J01XD | Imidazole derivatives | 2.41 | 70.12 | 14.30 | 8.75 | 5.53 | 4.29 | 105.40 |
| J01CA | Penicillins with extended spectrum | 7.43 | 5.98 | 35.23 | 7.96 | 5.97 | 27.04 | 89.60 |
| P01AB | Nitroimidazole derivatives | 4.35 | 12.09 | 12.42 | 13.14 | 11.66 | 10.96 | 64.62 |
| J01DD | Third-generation cephalosporins | 9.29 | 9.28 | 16.59 | 11.18 | 8.32 | 8.72 | 63.39 |
| J01CF | Beta-lactamase resistant penicillins | 4.32 | 3.86 | 31.77 | 4.26 | 4.54 | 6.91 | 55.66 |
| J01CE | Beta-lactamase sensitive penicillins | 2.55 | 2.45 | 2.04 | 2.18 | 2.96 | 7.87 | 20.04 |
| J01DB | First-generation cephalosporins | 1.98 | 1.24 | 0.07 | 0.88 | 2.07 | 2.73 | 8.97 |
| J01DH | Carbapenems | 0.37 | 0.36 | 0.53 | 0.60 | 0.34 | 0.77 | 2.97 |
| J01FF | Lincosamides | 0.05 | 0.08 | 0.07 | 0.12 | 0.50 | 1.97 | 2.79 |
| J01XA | Glycopeptide antibacterials | 0.00 | 0.06 | 0.01 |  | 0.13 | 0.20 | 0.40 |
| J01BA | Amphenicols | 0.01 | 0.08 | 0.10 | 0.02 | 0.01 | 0.06 | 0.28 |
| J01GB | Other aminoglycosides | 0.02 | 0.00 | 0.00 | 0.00 | 0.00 | 0.02 | 0.04 |
| Total |  | **81.37** | **169.79** | **279.62** | **107.29** | **272.65** | **288.12** | **1198.84** |

**Supplementary table 2: Antibiotic Consumption by class and Month (Total DDD/1000 inh. /day)**

| ATC Code | ATC Name | Apr-21 | May-21 | Jun-21 | Jul-21 | Aug-21 | Sep-21 | Total |
| --- | --- | --- | --- | --- | --- | --- | --- | --- |
| J01AA02 | doxycycline | 8.08 | 10.46 | 9.33 | 2.61 | 108.52 | 104.84 | 243.85 |
| J01CR02 | amoxicillin and beta-lactamase inhibitor | 10.00 | 10.38 | 108.28 | 9.72 | 8.49 | 17.08 | 163.95 |
| J01XE01 | nitrofurantoin | 3.43 | 5.74 | 3.99 | 3.98 | 58.93 | 55.38 | 131.45 |
| J01MA02 | ciprofloxacin | 8.20 | 20.55 | 24.21 | 11.92 | 21.92 | 22.87 | 109.66 |
| J01XD01 | metronidazole | 2.41 | 70.12 | 14.30 | 8.75 | 5.53 | 4.29 | 105.40 |
| J01FA09 | clarithromycin | 8.68 | 5.30 | 4.53 | 10.00 | 21.59 | 13.04 | 63.16 |
| J01FA10 | azithromycin | 8.18 | 8.79 | 15.52 | 19.27 | 9.32 | 1.04 | 62.11 |
| P01AB01 | metronidazole | 2.58 | 10.78 | 10.54 | 11.12 | 11.47 | 9.05 | 55.54 |
| J01CF02 | cloxacillin | 4.18 | 2.04 | 30.05 | 2.66 | 2.57 | 4.39 | 45.88 |
| J01CA01 | ampicillin | 4.17 | 2.04 | 30.05 | 2.66 | 2.57 | 4.39 | 45.87 |
| J01DD04 | ceftriaxone | 6.01 | 6.06 | 12.18 | 7.55 | 6.08 | 7.42 | 45.30 |
| J01CA04 | amoxicillin | 3.26 | 3.94 | 5.18 | 5.30 | 3.40 | 22.66 | 43.73 |
| J01CE02 | phenoxymethylpenicillin | 2.55 | 2.45 | 2.04 | 2.18 | 2.96 | 7.87 | 20.04 |
| J01DD63 | ceftriaxone and beta-lactamase inhibitor | 2.02 | 1.93 | 2.67 | 2.64 | 2.01 |  | 11.26 |
| J01CF05 | flucloxacillin | 0.13 | 1.82 | 1.72 | 1.60 | 1.97 | 2.53 | 9.78 |
| J01DB01 | cefalexin | 1.98 | 1.24 | 0.07 | 0.88 | 2.07 | 2.73 | 8.97 |
| P01AB02 | tinidazole | 1.34 | 0.94 | 1.44 | 1.51 | 0.00 | 1.35 | 6.58 |
| J01DD13 | cefpodoxime | 1.05 | 1.25 | 1.61 | 0.93 | 0.23 | 0.68 | 5.76 |
| J01MA06 | norfloxacin | 1.66 | 0.76 | 0.44 | 0.43 | 0.68 | 1.32 | 5.30 |
| J01FA01 | erythromycin | 0.35 | 2.21 | 0.19 | 0.26 | 1.18 | 0.98 | 5.18 |
| J01DH02 | meropenem | 0.37 | 0.36 | 0.53 | 0.60 | 0.34 | 0.77 | 2.97 |
| J01FF01 | clindamycin | 0.05 | 0.08 | 0.07 | 0.12 | 0.50 | 1.97 | 2.79 |
| P01AB07 | secnidazole | 0.28 | 0.24 | 0.23 | 0.28 | 0.00 | 0.39 | 1.41 |
| P01AB03 | ornidazole | 0.15 | 0.13 | 0.21 | 0.22 | 0.19 | 0.17 | 1.09 |
| J01DD52 | ceftazidime and beta-lactamase inhibitor |  |  |  |  |  | 0.62 | 0.62 |
| J01DD08 | cefixime | 0.21 | 0.04 | 0.13 | 0.05 | 0.00 | 0.00 | 0.44 |
| J01XA01 | vancomycin | 0.00 | 0.06 | 0.01 |  | 0.13 | 0.20 | 0.40 |
| J01BA01 | chloramphenicol | 0.01 | 0.08 | 0.10 | 0.02 | 0.01 | 0.06 | 0.28 |
| J01GB06 | amikacin | 0.02 | 0.00 | 0.00 | 0.00 | 0.00 | 0.02 | 0.04 |
| Total | | **81.37** | **169.79** | **279.62** | **107.29** | **272.65** | **288.12** | **1198.84** |
